# A Framework for Inferring and Analyzing Pharmacotherapy Treatment Patterns

**DOI:** 10.1101/2022.07.27.22277782

**Authors:** Everett Rush, Ozgur Ozmen, Minsu Kim, Erin Ortegon, Makoto Jones, Byung Park, Steven Pizer, Jodie Trafton, Lisa A. Brenner, Merry Ward, Jonathan Nebeker

## Abstract

**Objective:** To discover pharmacotherapy prescription patterns and their statistical associations with outcomes through a clinical pathway inference framework applied on real-world data.

**Materials and Methods:** We apply machine learning steps in our framework using a 2006 to 2020 cohort of veterans with major depressive disorder (MDD). Outpatient antidepressant pharmacy and emergency department visits, self-harm, and all-cause mortality data were extracted from the Department of Veterans Affairs Corporate Data Warehouse.

**Results:** Our MDD cohort consisted of 252,179 individuals. During the study period there were 98,417 cases of emergency department visits, 1,016 cases of self-harm, and 1,507 deaths from all causes. The top ten prescription patterns accounted for 69.3% of the data for individuals starting antidepressants at the fluoxetine equivalent of 20-39mg. Additionally, we found associations between outcomes and dosage change.

**Discussion:** For 252,179 Veterans who served in Iraq and Afghanistan with subsequent MDD noted in their electronic medical record, we documented and described the major pharmacotherapy prescription patterns implemented by VHA providers. Ten patterns accounted for almost 70% of the data. Associations between antidepressant usage and outcomes in observational data may be confounded. The low numbers of adverse events especially associated with all-cause mortality make our calculations imprecise. Furthermore, our outcomes are also the indications for both disease and treatment. Despite these limitations, we demonstrate the usefulness of our framework in providing operational insight into clinical practice, and our results underscore the need for increased monitoring during critical points of treatment.

## Introduction

Depression is the single largest contributor to disability worldwide ^1^. Major depressive disorder (MDD) is the most common form of depression ^2^. Between 2010 to 2019, the number of adults with at least one major depressive episode in the prior 12 months increased 25.2%, from 15.5 million to 19.4 million ^3,4^ While the cost of prescription drugs has decreased over time, the direct costs incurred by those with MDD rose 2.8% between 2010 and 2018 with medical services cost growing 18.1% and suicide-related costs growing 22.8% ^3^.

Veterans from Operation Iraqi Freedom (OIF), Operation Enduring Freedom (OEF), and Operation New Dawn (OND) have higher percentages of major depressive episodes than nonveterans of the same age ^5^. In response, the Department of Veterans Affairs (VA) has decreased suicides, improved drug monitoring programs, and increased population coverage by expanding mental health resources and by focusing existing resources on clinical practices that make the most difference ^6−10^. Existing reports and dashboards at VA cannot support policymakers with enough detailed insight into the full array of patient-level clinical treatment pathways to guide corrective action and resource provisioning efforts^11−15^.

This study presents a data-driven framework for providing explainable insights and finding critical decision points in the treatment of MDD with antidepressants. We present results of the framework using 15 years of pharmacy and administrative data on OEF/OIF veterans from VA’s Corporate Data Warehouse (CDW). Our approach uses state-of-the-art process mining techniques to describe prescription patterns and sequential rules mining to evaluate complex treatment patterns. Additionally, we propose a novel preprocessing technique that abstracts pharmacy data into both a trace suitable for process mining and a sequence database suitable for sequential rule mining.

## Methods

Our four-step framework takes pharmacy data and adverse events as input. 1) The preprocessing step normalizes all drug dosages to their fluoxetine equivalents, applies median smoothing to the prescribed daily dose, and abstracts the data into clinical actions. 2) We collect all the clinical actions for each depressive episode into a trace and apply agglomerative clustering to subdivide individuals based on their treatment pathway. Using these clusters, we formulate process inference as an optimization problem. 3) Additionally, we store the clinical actions for each depressive episode as a sequence database suitable for discovering statistically interesting rules correlated with adverse events. 4)We merge the discovered process models and association rules into a single process model. Figure 1 provides an overview of our framework.

**Figure 1.**
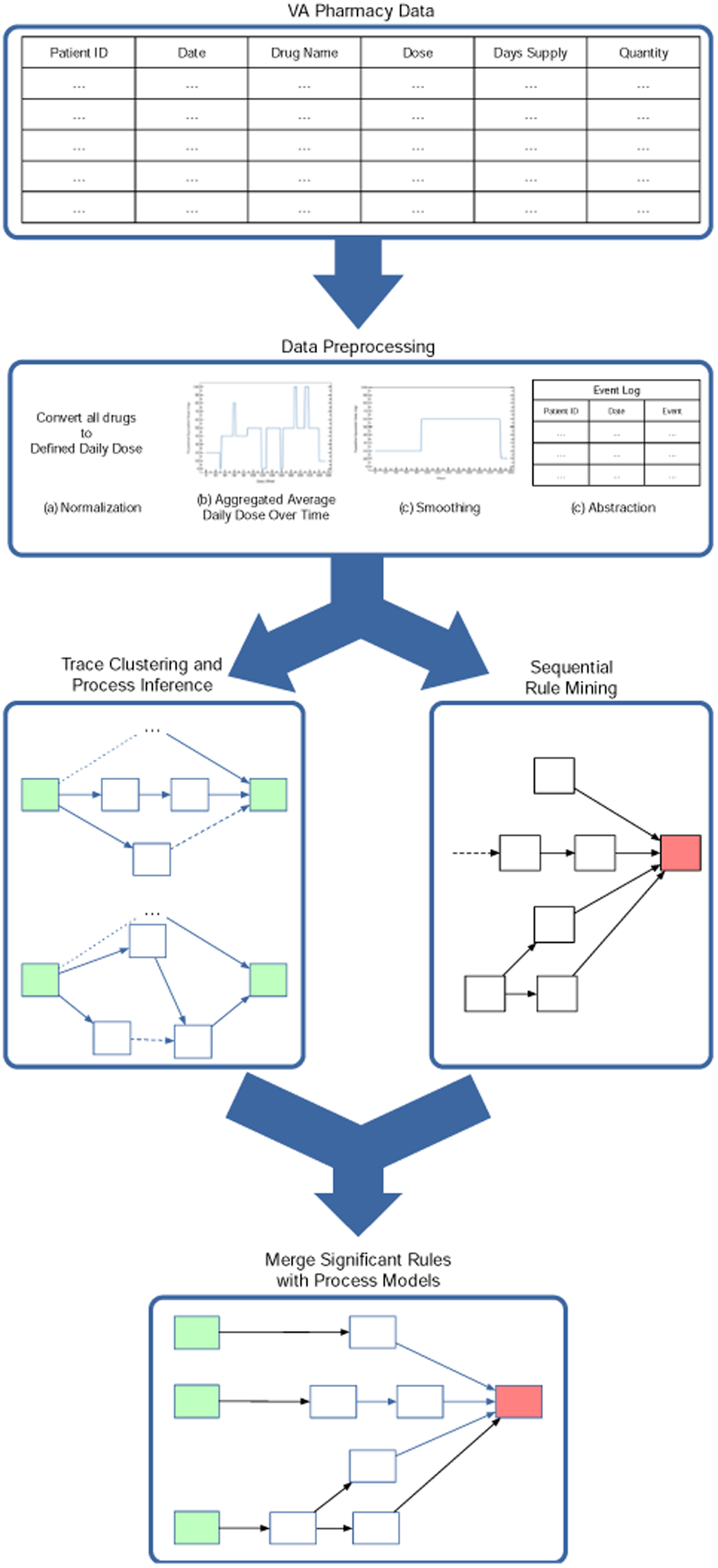
Overview of Inference and Analysis Framework

### Data Sources and Preprocessing

The CDW contains inpatient and outpatient pharmacy information for 12 million individuals aggregated from over 130 VA facilities across the United States. In this study, we used CDW data between January 1, 2006 and January 1, 2020 ^16^. We created a cohort of veterans that were diagnosed with MDD and served during OEF or OIF. This cohort contains younger veterans with fewer comorbid conditions that require MDD special considerations with prescribing antidepressants such as geriatric or hepatic populations. We considered a positive diagnosis of MDD as either one inpatient diagnosis or two outpatient diagnoses of an MDD ICD9 (296.20 -- 296.26) or ICD10 code (F32.0 -- F33.9). We used stop codes to determine emergency department visits, the self-directed violence classification system for self-harm, and the date of death from the CDW for all-cause mortality ^17^

We restricted the pharmacy data to the most common antidepressants prescribed at the VA in both inpatient and outpatient settings, i.e., sertraline, citalopram, fluoxetine, escitalopram, paroxetine, venlafaxine, duloxetine, trazodone, nefazone, and bupropion. These drugs comprise a majority of the antidepressant prescription fills for the MDD cohort.

We normalize all drug doses into their fluoxetine equivalent so that we can compare prescription patterns across drugs. While there are several conversion algorithms, we chose a combination of two methods: those of Hayasaka et. al. and the Defined Daily Dose (DDD) from the World Health Organization. Hayasaka et. al. use double-blind flexible dose trials to compute the optimal mean dose for each drug ^18,19^. On the other hand, the DDD is based on the drug’s product information sheet ^20,21^. Table 1 presents the drug equivalents used in this study. We adopt the dose equivalencies from Hayasaka et. al. where one exists and use the defined daily dose otherwise.

**Table 1.** For each drug in the study, this table provides equivalent milligrams between drugs.

| Drug | Defined Daily Dose | Hayasaka et. al. (2015) | Current Study |
| --- | --- | --- | --- |
| Sertraline | 50.0 | 49.3 | 49.3 |
| Citalopram | 20.0 | – | 20.0 |
| Fluoxetine | 20.0 | 20.0 | 20.0 |
| Escitalopram | 10.0 | 9.0 | 9.0 |
| Paroxetine | 20.0 | 17.0 | 17.0 |
| Venlafaxine | 100.0 | 74.7 | 74.7 |
| Duloxetine | 60.0 | – | 60.0 |
| Trazodone | 300.0 | 200.7 | 200.7 |
| Nefazodone | 400.0 | 267.6 | 267.6 |
| Bupropion | 300.0 | 174.25 | 174.25 |

The next preprocessing step approximates the milligrams of antidepressants that are consumed each day. For a given prescription, we use the fluoxetine equivalent dosage, days supply and quantity to determine the prescribed daily dose. Days with overlapping prescriptions are added together when a fill is released before the days supply of another prescription is over. Similar techniques are used in Coupland et. al. ^22,23^. We call the resulting metric the normalized prescribed daily dose.

The normalized prescribed daily dose can vary widely if a prescription is filled early or late. Gaps between prescriptions are treated as 0 mg that day and fills that are early are added on overlapping days. We smooth this curve by binning and using the median filter with a sliding window. Binning is a common approach used in previous studies ^22−24^ We binned average daily dose into five categories: <20mg, 20-39mg, 40-59mg, 60-80mg, and >80mg. Then we use a sliding window and the median function to smooth each data point. We calculated the size of the sliding window per person based on their average days supply.

Finally, we transformed each smoothed curve into a sequence of actions suitable for pathway inference. The action space mirrors actions from the Texas Medication Algorithms Project ^25^. Our action space *N* is start drug, increase dose, decrease dose, and continue at the current dosage. The start drug event occurs at the beginning of an episode when a drug is first started. The increase and decrease dose action corresponds to change from one bin to another with higher or lower dosage, respectively. The continue at current dose action specifies a duration of time over which the dosage stays constant.

### Trace Clustering

Antidepressant prescription patterns will vary based on the guidelines or algorithms being followed by the prescriber. There are a variety of MDD guidelines and algorithms available that can differ in significant ways ^26^. For example, there is disagreement on the second and third-line medications for MDD ^26,27,28^. Some guidelines suggest changing class while others suggest the use of tricyclic antidepressants. This variety in recommendations from guidelines leads to a multiplicity of guideline compliant prescription patterns.

Trace clustering is an effective technique to divide large complex logs into coherent groups that share similar characteristics ^29^. Many approaches build process models and cluster the process models or cluster individual traces directly. Current approaches work directly with traces and represent each trace as a vector that can be easily clustered ^30−33^. We prefer to represent each trace *t* as an matrix *T*^*nxn*^ where *n* is the cardinality of action space *N* and *T*_*ij*_ = 1 if action *n*_*j*_ directly follows action *n*_*i*_ in *t*. We use the Frobenius norm to calculate the distance || *A* − *B* ||_*F*_ between each pair of matrices *A* and *B*. Then we use agglomerative hierarchical clustering to cluster similar traces ^29^.

### Process Inference

The collection of all actions for a single medication episode is a trace. The collection of all traces across the entire cohort is called an event log. Given an event log, we wanted to find the process model *PsM* with bounded complexity that performs best under a given metric. A *PsM* is defined by a set of nodes and edges such that *PsM* = (*N, E*) where | *E* | is fixed. For *n* ∈ *N, n* is an action from the event log. For a given edge (*n*_*i*_, *n*_*j*_) ∈ *E*, the edge denotes that *n*_*i*_ occurs directly before *n*_*j*_ in the *PsM*.

The replayability game scores each *PsM* based on a given replayability function *R* over a given log *L*. The replayability score of an individual trace σ ∈ *L* using *PsM* is *R(PsM*, σ). Prodel et. al. introduced eight replayability functions with varying properties ^34^ The *R*^*4*^ function gives the percentage of events replayed with a penalty for skipping events. The *R*^*4*^ function is defined as

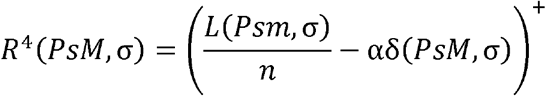

where *L(PsM*, σ) denotes the cardinality of the longest subsequence of trace σ that can be replayed using *PsM*, α is a constant, and δ is a binary indicator variable activated when an event is skipped. The *R*^*4*^ score is then averaged across all traces in the event log.

We used a heuristic algorithm to search for a *PsM* that scored highest with respect to the *R*^*4*^ function. We utilized a Python implementation of Tabu Search where the local moves are based on edge frequency.

### Sequential Rule Mining

Sequential Rule Mining is a data mining technique used to extract sequential patterns from a sequence database. A sequential rule has the form *X* ⇒ *Y* and is read if a sequence of events *X* occurs then another sequence of events *Y* is likely to occur. Two definitions for sequential rule mining exist, and we adopt the same definition of sequential rules as Fournier-Viger et. al. ^35−37^ Formally, there is a set of sequences S = {s_1_, s_2_,…, s_n_} and a set of items I = {i_1_ i_2_,…, i_m_}. A sequence s_j_ is an ordered list of itemsets S_j_ = {I_1_, I_2_,…, I_t_}. For two unordered itemsets X, Y ⊆ I, a sequential rule states that if the items of *X* occur in a sequence, then the items in *Y* will occur afterward in the same sequence.

We use different statistical measures to assess the interestingness of the sequential association rules. For each sequential association rule *X* ⇒ *Y* we capture support, confidence, and odds ratio. Support is defined as *P(X, Y*) Confidence is defined as P(Y|X). Since adverse events happen with very small frequency, support and confidence do not describe the sequential rules well. To remedy this problem, we also report the odds ratio as

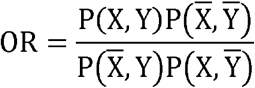

previously defined in Tan et. al. ^38^. A high odds ratio metric denotes that a sequence containing X is more likely to have *Y* than sequences without X.

### Merge Rules with Clinical Context

The sequential rules themselves do not provide enough context around mined associations. For example, an association that happens frequently after months of treatment is different than one found at the very beginning of treatment. This step uses a bow-tie analysis where we consider events within a specific time frame of a rule of interest to generate enough clinical context to understand the association rule ^13^.

First, we create a log with the traces associated with a sequential rule of interest. We truncate each trace to only consider events within a uniform amount of time of the adverse event. Using the time filtered log, we perform process inference. We visualize the clinical context using a custom pathway visualization tool on our Github page ^39^.

## Results

The OEF/OIF MDD cohort has 458,352 individuals with an average age of 41.7 years. We found antidepressants in VA pharmacy records for 252,179 out of the 458,352 individuals from the OEF/OIF MDD cohort (55%). We applied our data preprocessing methodology and created 288,344 antidepressant medication traces. During treatment with antidepressants there are 98,417 cases of emergency department visits, 3,928 cases of self-harm, and 1,507 deaths from all causes. Table 2 provides a summary of the data.

**Table 2.** Total number of traces and the number of individuals with adverse events of given types.

| Outcomes |  |
| --- | --- |
| Total Traces | 288,344 |
| ED Visits | 98,417 |
| Self-Harm | 1,016 |
| All-cause Mortality | 1,507 |
| Demographics |  |
| Race |  |
| White Nonhispanic | 274,879 |
| Black Nonhispanic | 84,233 |
| Hispanic | 57,529 |
| Other | 41,056 |
| Gender |  |
| Female | 388,006 |
| Male | 69,691 |

### Trace Clustering and Process Inference

After trace clustering and process inference, we had 3,607 different clusters each with their own process model. We achieve an average *R*^4^ score of 0.891 across all traces.

The process models from the 10 largest clusters in each bin completely replay 69.3% of all traces which we present in Figure 2 (top 10 process models for traces that start at 20-39mg). These 10 process models completely replay 90,297 of the 130,207 traces. Figure 2 provides a visualization of each process model along with the number of traces completely replayed on the process model. The images are sorted in descending order based on the number of traces that can be completely replayed. For example, Figure 2a shows the most common prescription pattern. Figure 2a shows that the normalized prescribed daily dose stays in the 20-39mg dose range for 4 or more months with some discontinuing at a lower dose or staying on the lower dose long-term.

**Figure 2.**
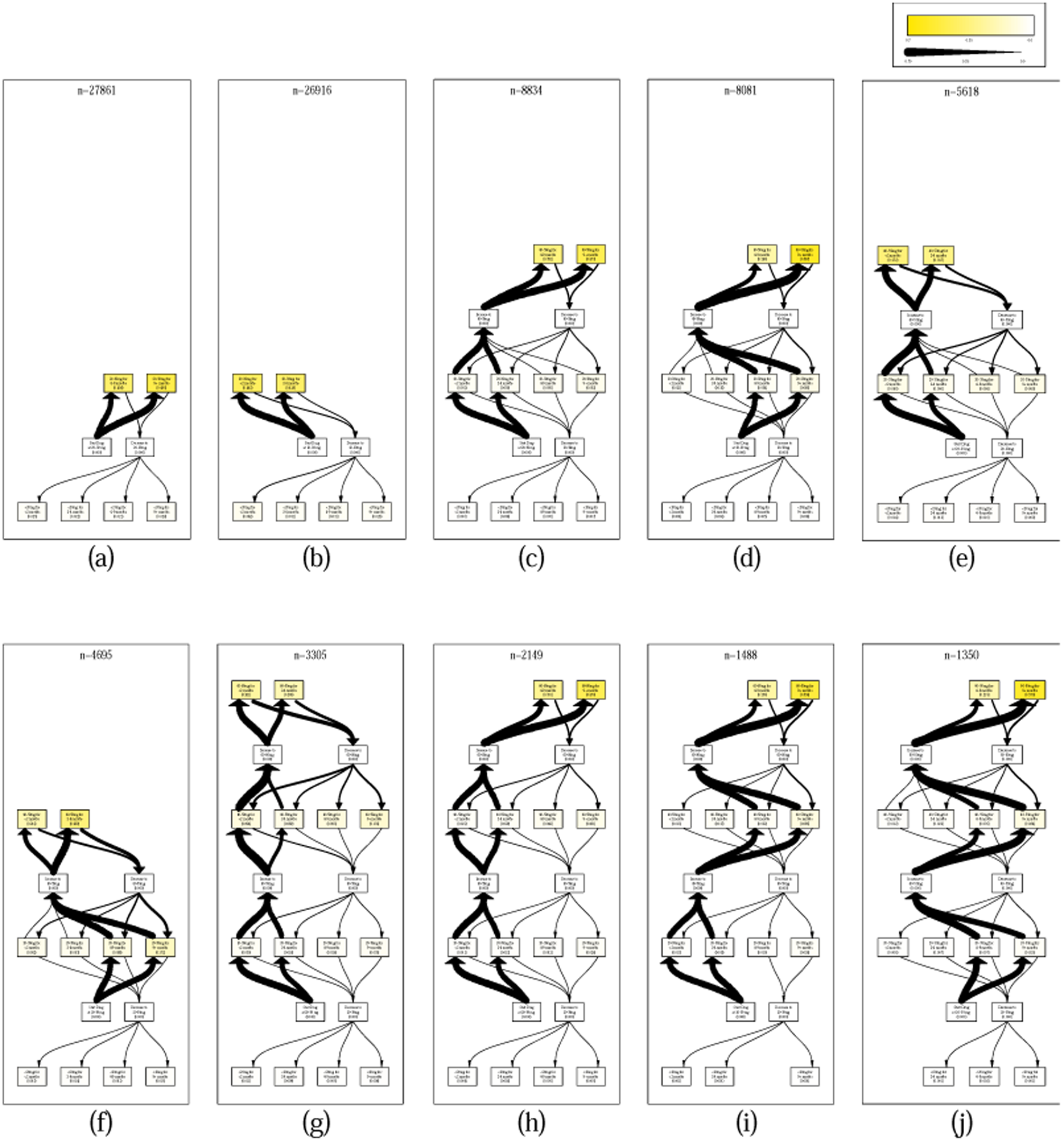
This figure shows the top 10 process models that start at 20-39mg. The value in each box represents the percentage of people whose treatment stopped with this activity. The thickness of arrows is a normalized edge weight with edge frequency divided by *n*.

### Sequential Rule Mining

We created sequences by dividing up the smoothed prescribed daily dose curve into two-week increments. We choose two week increments by looking at the distribution of the amount of time from start or dosage change to adverse event and two weeks was a good cut-off point. Additionally, this enables us to compare our results with existing literature that suggests adverse drug reactions occur within the first two weeks after dosage change ^40^. Then we inserted the first adverse event into each sequence. We considered emergency department visits, documented selfharm, and all-cause mortality. After mining sequential association rules using window size of 1, 2, and 3, we kept rules with support greater than 10 and an odds ratio greater than 2.0.

Table 3 presents the top 10 sequential association rules for emergency department visits and selfharm. Length one rules were mined using a window size of 1, and length two rules were mined with window size of 2. There were no rules that met the support and odds ratio thresholds. Each section is sorted by increasing odds ratio. There were no significant length two rules found for self-harm and all-cause mortality. Note that these association metrics do not claim that there is a casual relationship between outcome and treatment

**Table 3.**
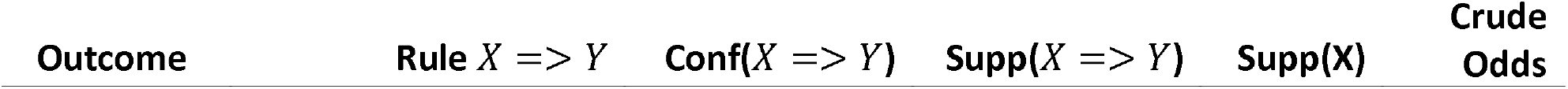
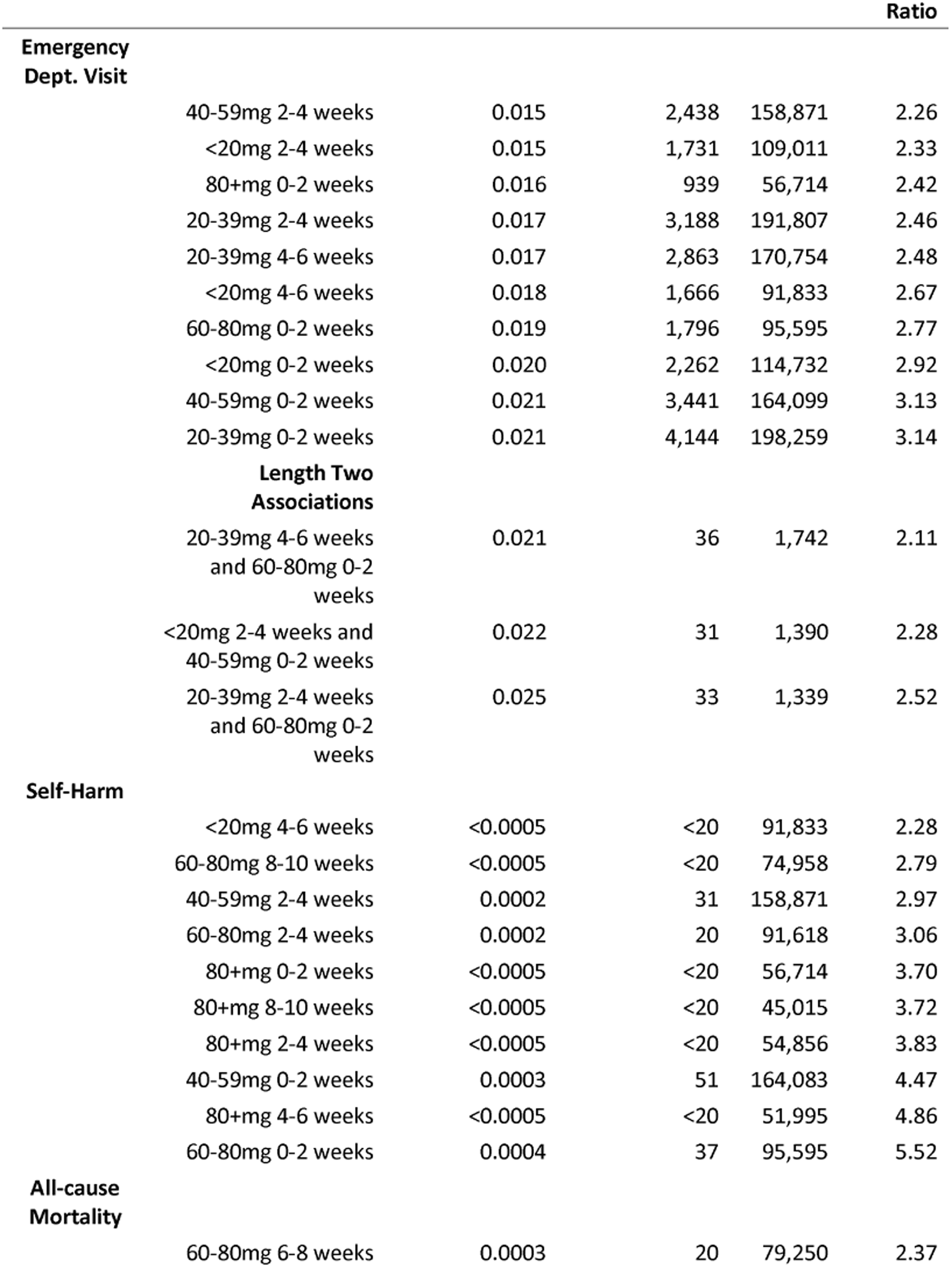
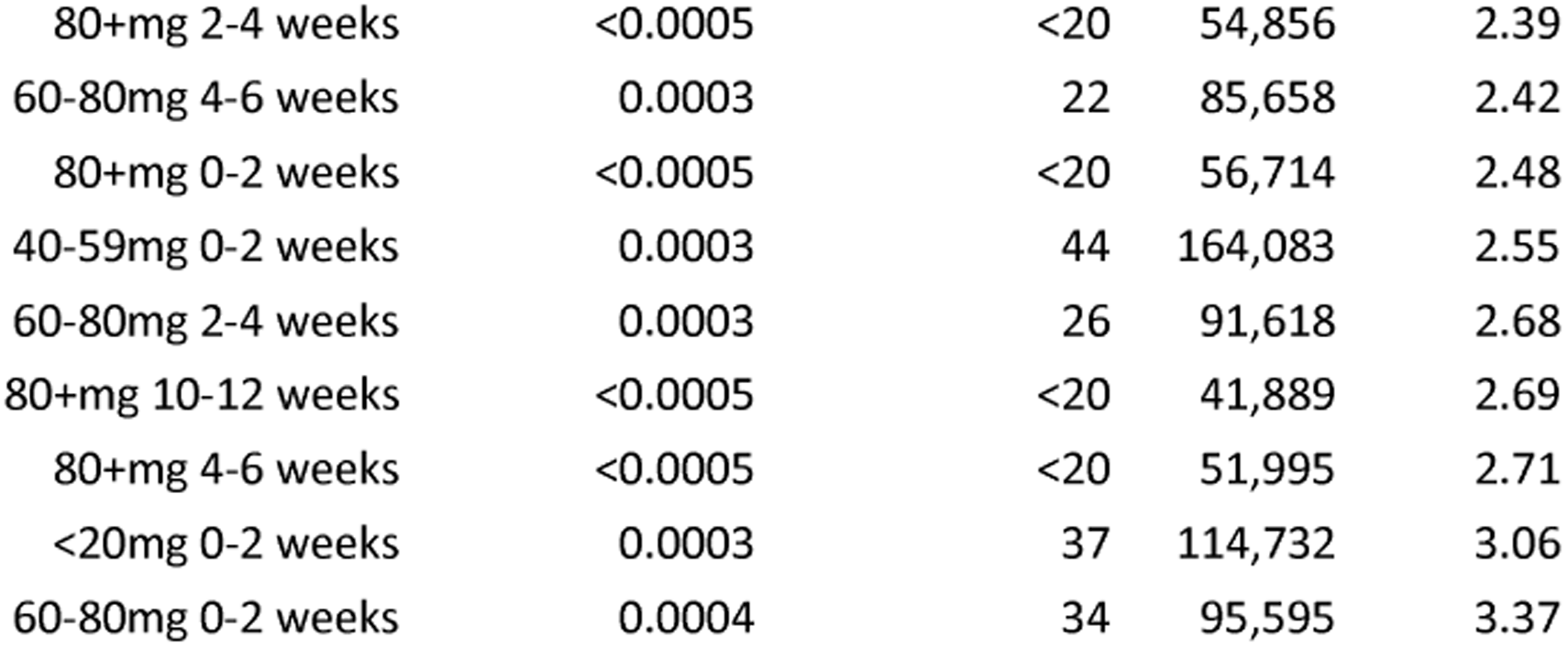
This table provides sequential association rule metrics for associations between time duration, dosage, and outcomes.

### Merge Rules with Clinical Context

Figure 3 shows the clinical context for the sequential association rule (80 + mg for 0 − 2 weeks ⇒ *self* − *harm*). This rule is interesting because it shows the recent history of individuals on a high dose of antidepressants (OR=3.70). The confidence metric for this rule shows 0.03% of individuals (n=17) that go above 80mg (n=56,716) have their first documented occurrence of self-harm within 0-2 weeks after increasing dosage.

**Figure 3.**
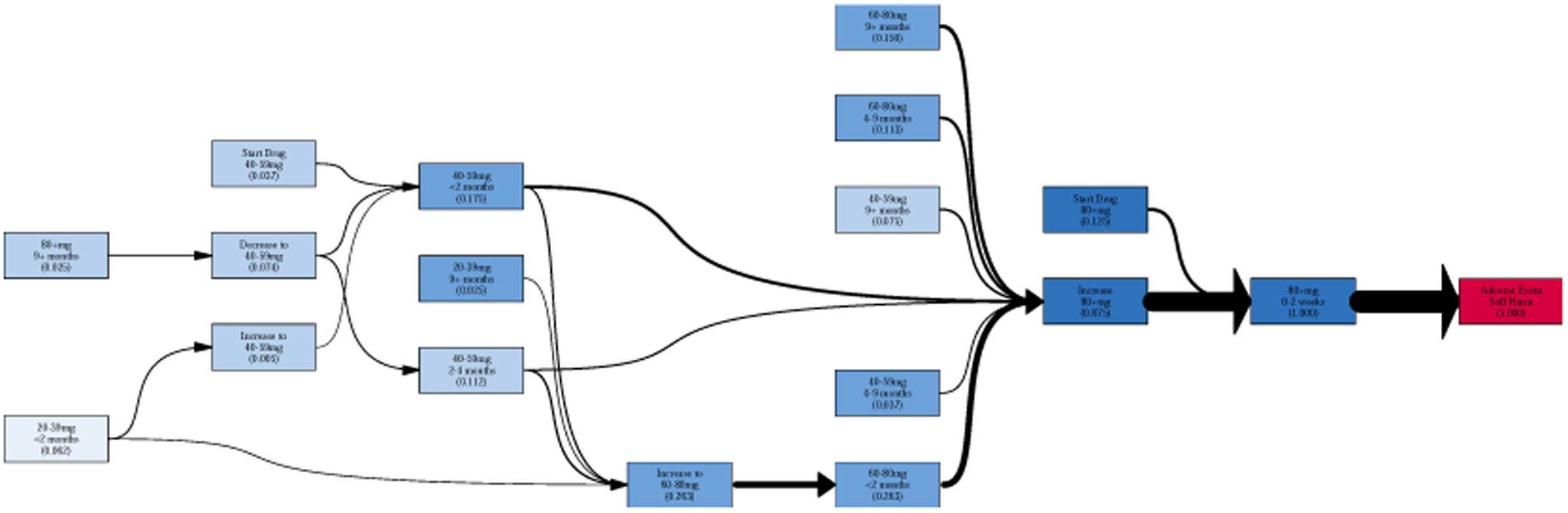
This image shows the clinical context for the sequential association rule (80 + *mg* 0 − 2 *weeks* ⇒ self − *harm*). The activities (boxes) are colored by dosage. The adverse event is highlighted in red. The thickness of each arrow represents the normalized edge frequency.

## Discussion

We demonstrated that it is possible to describe a majority of antidepressant prescription patterns with a small number of process models. That is, 10 models accounted for almost 70% prescription patterns. Furthermore, we derive new insights into critical points during MDD treatment. These insights can help inform drug monitoring efforts and suicidal behavior risk.

We found that 55.0% of the entire OEF/OIF MDD cohort took SSRIs or SNRIs at some point during their depressive episode. This is in line with national trends ^41^. Most traces start initial dosing at either <20 mg (n=76,925) or 20-39 mg (n=130,207). The number of people at a suboptimal dose is much higher than the national average or 16% reported by Lou et. al. Even after conducting a separate analysis using the FDA therapeutic dose definitions, the number of individuals starting at suboptimal doses remains high.

The replayability score of 0.891 shows that the discovered process models are able to replay more than 89% of the trace data. Our models were unable to incorporate low frequency events such as drastic dosage changes of +60mg or -60 mg. While we can include those transition in the process models, the models are less interpretable and tend to look more like spaghetti. So, we choose to restrict the number of edges in each process model in favor of more interpretable results with lower replayability score.

Figures 2a-d account for a majority of the people that starts at 20-39mg. The most frequent prescription pattern is for people to stay in the 20-39mg range for more than 4 months. Figure 2b shows individuals discontinuing treatment in <2 months (n=11,924) or between 2-4 months (n=11,251). Figure 2c and d show almost equal numbers of people going up to the 40-59mg range for 4 or more months. Figure 2c spends <4 months at the 20-39mg range whereas Figure 2d spends 4 or more months at 20-39mg. Figures 2c and 2h show a fast titration schedule going to higher doses of antidepressants with continued treatment for 4 or more months at the highest dosage. Figures 2d, 2i and 2j show a slow titration schedule that can happen over 4 or more months.

The treatment patterns for those individuals that start at higher doses are more varied than those that start at lower doses. The top 10 process models in the 20-39mg bin replay 69.3% of traces that start at 20-39mg. In contrast, the top 10 models in the 40-59mg range and 60-80mg range only replay 55.9% of traces and 50.5% of traces respectively.

The rules from Table 3 do not infer causation between treatment and outcome, but instead the rules capture the temporal order that events happen together. While Table 3 shows many rules occur in the first four weeks after starting treatment or a dosage change, we speculate that prescribers notice that treatment isn’t working, and they are trying to adjust the dosage. We also note that there is a possibility of capturing adverse drug reactions as well. Stübner et. al. report that 93% of the suicidal adverse events from the European drug surveillance program occur within the first weeks of starting antidepressant medication or increasing dose ^39^.

Emergency department (ED) visits were the most frequent adverse events. The top four rules in this category all occur within the first two weeks of starting treatment or changing doses. There are only two rules identifying an association beyond 4 weeks. Interestingly, the length two associations all involve an increase in dose greater than 20mg. The self-harm outcome has many of the same rules as the emergency department visit. The top five rules all occur in the first two weeks after starting treatment or changing dose. Two rules occur after 4 weeks. The length two associations all occur in the first two weeks after a dosage change at high milligram. Death from all causes is the lowest frequency outcome in the study with 1,507 deaths or approximately 0.6% of the cohort. The support for the top 10 rules sums to 15.8% of total number of deaths.

The sequential association rules must also be viewed in their clinical context. Figure 3 shows the clinical context for self-harm that happens within the first two weeks after going above 80mg. The rule is read as within 0-2 weeks after going above 80mg the first documented care of self-harm happens with OR=3.70. The average amount of time until the first documented self-harm event is 72.3 weeks. 11.75% of the traces that the rule applies to start at 80+ mg and within 2 weeks have a documented self-harm event. The other 76.5% were on a lower dose and increased to 80+ mg. For those that did not start at 80+mg, they either came from the 40-59mg range or the 60-80mg range. These results underscore the need for careful monitoring during these time periods.

This study has some limitations. First, observational data has sources of both biases and confounders. Indication bias is one large source of error. Indication bias arises because the outcomes of the study are indications of MDD and side-effects of treatment. Furthermore, we do not account for severity of depression or individuals with pre-existing conditions related to our outcomes. Next, the actual consumption of antidepressants is different than the prescribed daily dose. Consequently, we are unable to account for individuals that stock-pile medications and use them later.

Due to the limitations with our approach and data, we don’t see this approach as a way to inform or influence clinical practice. We present this approach as a documentation and data exploration step that can provide a basis for further efforts into medication management. Additionally, there is potential to use the results from this framework in medication monitoring efforts.

## Conclusion

This study presented a data-driven framework for inferring pharmacotherapy treatment patterns and showcases a proof-of-concept study on pharmacotherapy prescription patterns used to treat MDD. Using pharmacy records for 252,179 individuals from an OEF/OIF cohort with MDD we documented and described the major pharmacotherapy prescription patterns implemented in the VA. We. We also added three outcomes to enable an association study between outcomes, drug dosage and treatment duration. We presented sequential association rules that link drug dosage and duration with outcomes. Then we presented a method for placing each rule in their clinical context for further investigation. Our results underscore the need for increased monitoring at certain points in pharmacotherapy treatment of MDD.

Our initial findings show that this is a promising approach for inferring and analyzing prescription patterns. We don’t claim any causal relationship for our association rules. Future work is needed to perform a causal analysis between medications prescription patterns and outcomes.

## Data Availability

All data produced in the present study are available upon reasonable request to the authors

## Acknowledgments

This work is sponsored by the US Department of Veterans Affairs. This research used resources from the Knowledge Discovery Infrastructure at Oak Ridge National Laboratory, which is supported by the DOE Office of Science under contract DE-AC05-00OR22725.

## Notes

### Competing Interest Statement

The authors have declared no competing interest.

### Funding Statement

This study was funded by Veterans Affairs.

### Author Declarations

Oak Ridge Sitewide Institutional Review Board (IRB#: IRB00000547) determines a work using the data applied in this study to be EXEMPT based on the applicable federal regulations, including DOE O443.1C and has been approved. Approval date is 4/11/2022 and expiration date is 4/10/2023.

